# PREVALENCE OF POST STROKE DEPRESSION AND ANXIETY AMONG PATIENTS ADMITTED IN KHYBER TEACHING HOSPITAL PESHAWAR

**DOI:** 10.1101/2023.04.10.23288355

**Authors:** Sayed Sajid Hussain, Zaib Ullah, Mahmood Bacha, Sheikh Usman Khalil

**Affiliations:** Sarhad University of Science and IT, Peshawar; Sarhad Institute of Allied Health Sciences, SUIT, Peshawar

**Keywords:** Stroke, Post stroke Depression, Post stroke Anxiety

## Abstract

**INTRODUCTION:** Stroke is a life-threatening medical condition that can result in lifelong brain damage, complications, and death. It is the second leading cause of death worldwide and could soon overtake as the biggest cause of death globally. This can be produced by ischemia from a thrombosis or embolism, or a hemorrhage. As a result, the brain’s damaged area is unable to operate, causing one or more limbs on one side of the body to become immobile, inability to recognize or produce speech, or seeing only one side of the visual field.

**OBJECTIVE OF THE RESEARCH:** To determine the prevalence of Post Stroke Depression and anxiety among stroke patients admitted in Khyber teaching hospital Peshawar

**METHODOLOGY:** A descriptive cross-sectional study was carried out to determine the prevalence of PSD and anxiety among stroke patients admitted at Khyber teaching hospital Peshawar. Data was collected through a questionnaire named HADs scale from participants. The sample size calculated for the research study was 109. The convenience sampling technique was used in the current study.

**RESULTS:** The findings of the study provide a baseline of information about the prevalence of Post Stroke Depression (PSD) and anxiety among stroke patients admitted in Khyber teaching hospital Peshawar. The prevalence of depression was 86(63.3%) and anxiety was 91(83.5%).

**CONCLUSION:** In current study we had concluded a high prevalence of post stroke depression and anxiety ranging from 65-74 years of age in Khyber teaching hospital Peshawar. So, we concluded that females suffering from stroke have more chances of post stroke depression.

## INTRODUCTION

### STROKE

Stroke is a life-threatening medical condition that can result in lifelong brain damage, complications, and death. It is the second leading cause of death worldwide and could soon overtake as the biggest cause of death globally. A stroke is the loss of brain functions that occurs rapidly as a result of a disruption in the blood vessels providing blood to the brain. This can be produced by ischemia from a thrombosis or embolism, or a hemorrhage. As a result, the brain’s damaged area is unable to operate, causing one or more limbs on one side of the body to become immobile, inability to recognize or produce speech, or seeing only one side of the visual field.(Marwat, Usman, & Hussain, 2009, 2017) Martin Roth (Roth, 2010) was the first to investigate the link between depression and stroke. Folstein later found that depression was more common in stroke patients than in patients with similar levels of motor impairment due to orthopedic issues. (Folstein, Maiberger, & McHugh, 2015) Several studies have discovered a link between depression and quality of life in stroke survivors. (Gbiri & Akinpelu, 2012) In fact, past research has shown that the prevalence of PSD is impacted by the patient’s choice, socioeconomic status, and risk factors. Unfortunately, PSD is frequently under diagnosed and underreported, in part because cognitive impairments following a stroke can muddle depression symptoms and make diagnosis difficult. Despite the growing interest in the socioeconomic implications of chronic medical illnesses in developing nations, little is known about PSD in Egypt. To address this issue, we conducted a study in Qena University Hospital to determine the prevalence of PSD, associated risk factors, and impact on quality of life (QOL) among stroke patients.(Cai, Mueller, Li, Shen, & Stewart, 2019).

### ANXIETY

Anxiety is a widespread and unfocused feeling of uneasiness and anxiety caused by an overreaction to a circumstance that is only subjectively seen as dangerous.(Chang, 2020). It comprises negative subjective sensations of dread about upcoming events. (Davison, 2017). Muscle tension, restlessness, weariness, inability to catch one’s breath, abdominal tightness, and attention issues are all common symptoms. Anxiety is closely related to fear, which is a reaction to a real or perceived immediate threat; anxiety, on the other hand, is a response to the anticipation of a future threat, including dread. (American Psychiatric Association, 2016).

### TYPES OF STROKE ISCHEMIC STROKE

An ischemic stroke occurs when blood supply to a portion of the brain is cut off, causing brain tissue in that area to malfunction. There are four possible causes for this: Thrombosis is a type of thrombosis that occurs when (obstruction of a blood vessel by a blood clot forming locally), Embolism is a type of blood clot that (obstruction due to an embolus from elsewhere in the body). (Ga, 2018).

### HEMORRHAGIC STROKE

The underlying pathology of hemorrhagic strokes is classified. Hypertensive hemorrhage, ruptured aneurysms, ruptured AV fistulas, transformation of earlier ischemic infarction, and drug-induced bleeding are some of the causes of hemorrhagic stroke. (Kasper et al., 2015) They cause tissue harm by compressing tissue as a result of a growing hematoma or hematomas. Furthermore, the pressure may cause a loss of blood supply to the damaged tissue, resulting in an infraction, and the blood discharged by a brain hemorrhage appears to have direct harmful effects on brain tissue and the vasculature. (Feldman, Cornblath, Porter, Dworkin, & Scherer, 2015) Inflammation plays a role in subsequent brain injury following a bleed. (Wang, 2017).

### POST STROKE DEPRESSION

Depression affects around a third of stroke survivors at any given time and is linked to poor functional outcomes(Hackett & Pickles, 2014) and increased mortality. (Bartoli et al., 2013). PSD is also a strong predictor of poor functional outcomes following a stroke. PSD is linked to poorer functional and cognitive recovery, increased constraints on daily life activities as well as social and interpersonal activities, lower quality of life, and higher mortality rates (up to ten times higher than in patients without PSD). (Angelelli et al., 2014; Pohjasvaara, Vataja, Leppävuori, Kaste, & Erkinjuntti, 2016). Furthermore, we know that there are a number of risk factors for developing PSD, including a severe motor deficiency, a higher level of impairment, and a weakened social support network. The early identification of these factors allows for the implementation of preventative and treatment initiatives. (F. Carod-Artal, 2016).

### EPDEMIOLOGY

Depression is frequent after a stroke, affecting roughly one-third of stroke survivors at any given time (compared to 5%–13% of persons without a stroke), with a cumulative incidence of 55 percent.(Hasin, Goodwin, Stinson, & Grant, 2015; Kessler, Sonnega, Bromet, Hughes, & Nelson, 2015) Estimates of lifetime prevalence range from 3% in Japan to 17 percent in the United States. According to epidemiological research, the Middle East, North Africa, South Asia, and the United States of America have greater incidence of depression than other countries. (Ferrari et al., 2013) In most of the ten nations analyzed, the percentage of persons who will experience depression at some point in their life falls between 8 and 12 percent.(Andrade et al., 2013; Dummer et al., 2018).

### SYMPTOMS OF PSD

PSD can develop quickly after a cerebrovascular incident; however it usually appears within the first several months after a stroke. (F. Carod-Artal, 2016; Dafer, Rao, Shareef, & Sharma, 2018) Although depressive symptoms are often identical, there are notable variations between PSD sufferers and others who have depression but no neurological disorder. Patients with late-onset PSD, particularly those with right-hemispheric damage, are less likely to have dysphoric depression show more social disengagement and fewer agitation symptoms. (Lökk & Delbari, 2010; Pohjasvaara, Vataja, Leppävuori, Kaste, & Erkinjuntti, 2015) As a result, it’s critical to conduct a differential diagnosis to rule out other cognitive-behavioral diseases that present with mood and affective changes following a stroke, as their symptoms may be similar to PSD’s. (Artal, 2016).

## MATERIAL AND METHODS

After approval of Advance Studies and Research Board (ASRB), graduate and more likely to exhibit vegetative signs and symptoms. (Lökk & Delbari, 2015; Paradiso, Vaidya, Tranel, Kosier, & Robinson, 2018). When compared to depressive patients with other disorders, older stroke patients with depression committee and Ethical board of Sarhad University of Science and Information Technology the data was collected.

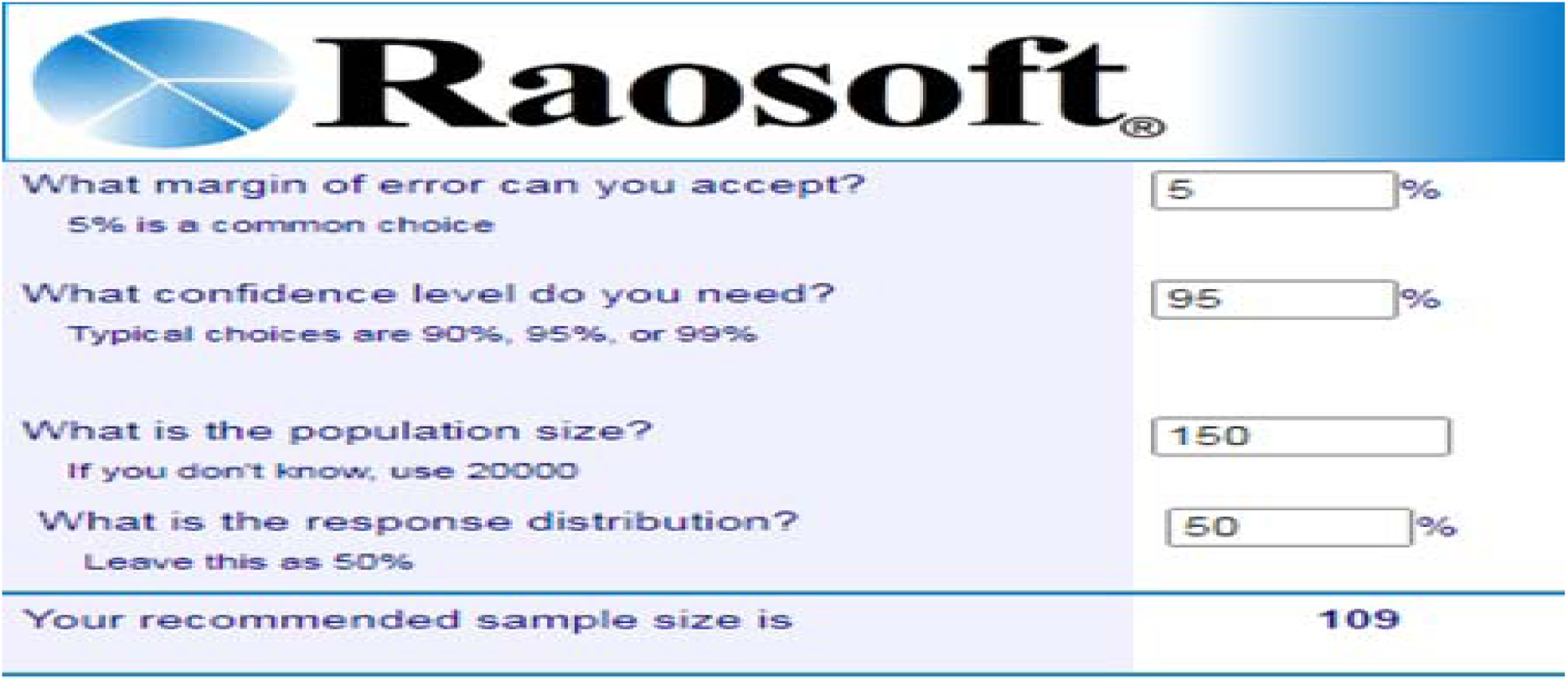

### STUDY DESIGN

This study is descriptive cross-sectional study conducted on post-stroke patients suffering from post-stroke depression and anxiety admitted in Khyber teaching hospital Peshawar.

### Study Population

All the patients’ males and females, above 18 years suffering from Post Stroke Depression and anxiety admitted in hospital were the population for this study.

### STUDY DURATION

The time frame of the research was six months.

### SAMPLE SIZE

Sample size for current research study was calculated by Raosoft and sample size was 109.

### SAMPLING TECHNIQUE

We use convenience sampling technique in current study. It is a non-probability technique.

### INCLUSION CRITERIA

1. Acute post-stroke patients admitted in hospital.
2. Age 18 years or older were included.
3. Those patients who were oriented with persons, time, and place; and had no problems in communication.
4. Both male and females are included.
5. Those that is willing to participate in the study.

### EXCLUSION CRITERIA

1. Individuals who were unable to communicate due to a disturbed level of consciousness.
2. Comatose patients were excluded.
3. Patients with Other Comorbidities were excluded.

### DATA COLLECTION PROCEDURE

After approval of the topic from the research committee of our institute (Advance Studies and Research Board (ASRB), graduate committee and Ethical board of Sarhad University of Science and Information, Peshawar), a cross-sectional survey were conducted to determine the prevalence of post stroke depression and anxiety at Khyber teaching hospital Peshawar. Every aspect of the ethical issue related to the current research survey was thoroughly reviewed by the responsible member of the research committee of the suit University System, Department of Physical Therapy and Rehabilitation. Each feature of the research study were a promising assurance about the confidentiality and privacy of the participants. The consent form was taken from all the participants of the study. Data were collected through Hospital Anxiety and Depression Scale (HADS) which measure the severity of depressive symptoms according to the subject score as follow: 0-7=normal, 8-10=borderline abnormal, 11-21=abnormal case.

## RESULTS

### THE AGE OF THE PARTICIPANT

| Age | Prevalence | Percent | Cumulative Percent |
| --- | --- | --- | --- |
| 65-74 | 97 | 89.0 | 89.0 |
| 75-84 | 6 | 5.5 | 94.5 |
| greater than 84 | 5 | 4.6 | 99.1 |
| Total | 109 | 100.0 |  |

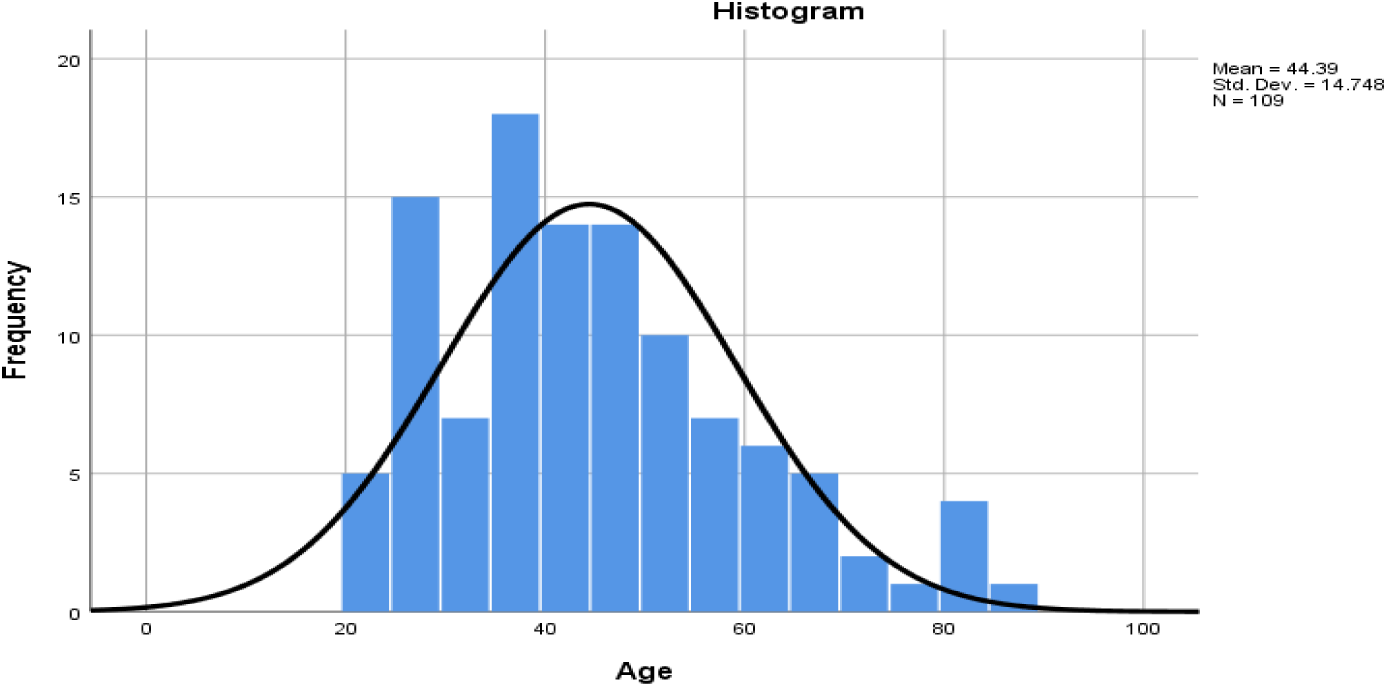

The above table and figure show the age of the participants in years. The participant’s age ranging from 65-74 years is 97(89.0%) followed by 75-84 years 6(5.5%) and greater than 84 years 4.6(4.6%).

### I FEEL TENSE AND WOUND UP

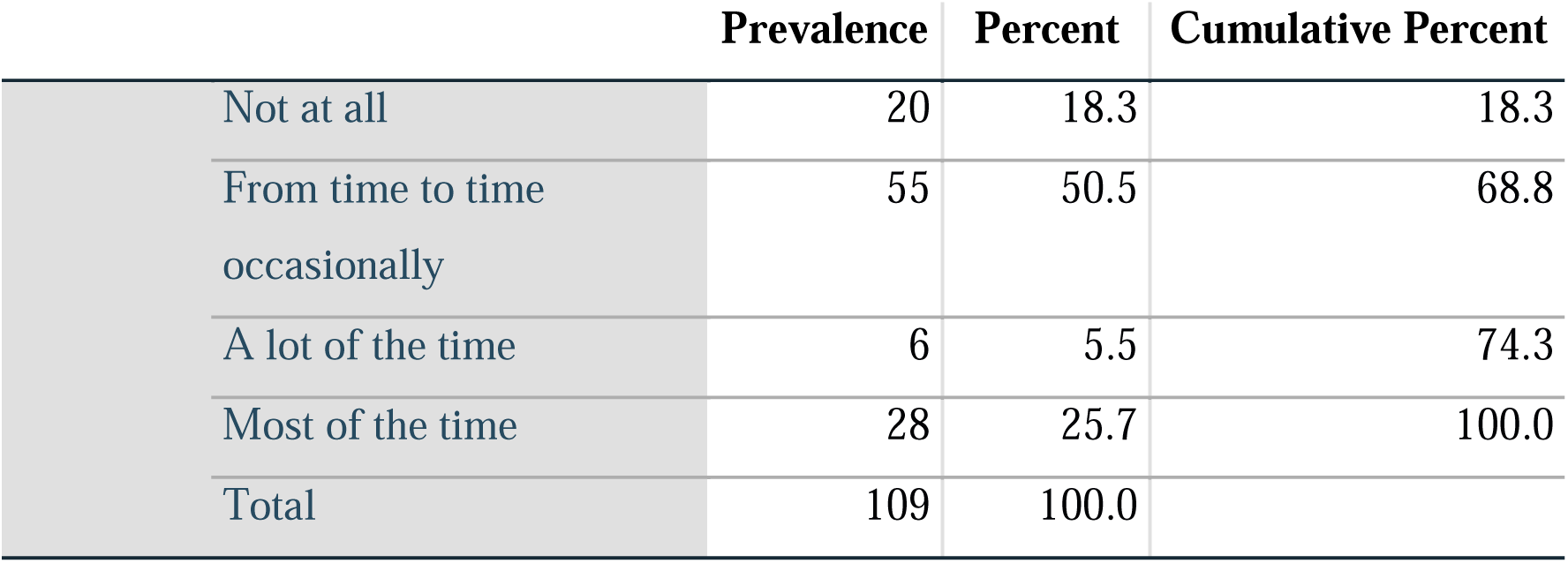

|  | Prevalence | Percent | Cumulative Percent |
| --- | --- | --- | --- |
| Not at all | 20 | 18.3 | 18.3 |
| From time to time occasionally | 55 | 50.5 | 68.8 |
| A lot of the time | 6 | 5.5 | 74.3 |
| Most of the time | 28 | 25.7 | 100.0 |
| Total | 109 | 100.0 |  |

The above table shows that the participants feel tense and wound up from time to time occasionally 55(50.5%) followed by most of the time 28(25.7%), Not at all 20(18.3%) and A lot of the time have 6(5.5%).

### GET SORT OF FRIGHTENED FEELING AS IF SOMETHING AWFUL IS ABOUT TO HAPPEN

|  | Prevalence | Percent | Cumulative Percent |
| --- | --- | --- | --- |
| Not at all | 20 | 18.3 | 18.3 |
| A little, but it does not worry me | 5 | 4.6 | 22.9 |
| Yes, but not too badly | 69 | 63.3 | 86.2 |
| Very definitely and quite badly | 15 | 13.8 | 100.0 |
| Total | 109 | 100.0 |  |

The above table shows that the participants get sort of frightened feeling as if something awful is about to happen is yes, but not too badly 69(63.3%), followed by not at all 20(18.3%), very definitely and quite badly 15(13.8%) and a little, but it does not worry me 5(4.6%).

### I HAVE LOST INTEREST IN MY APPEARANCE

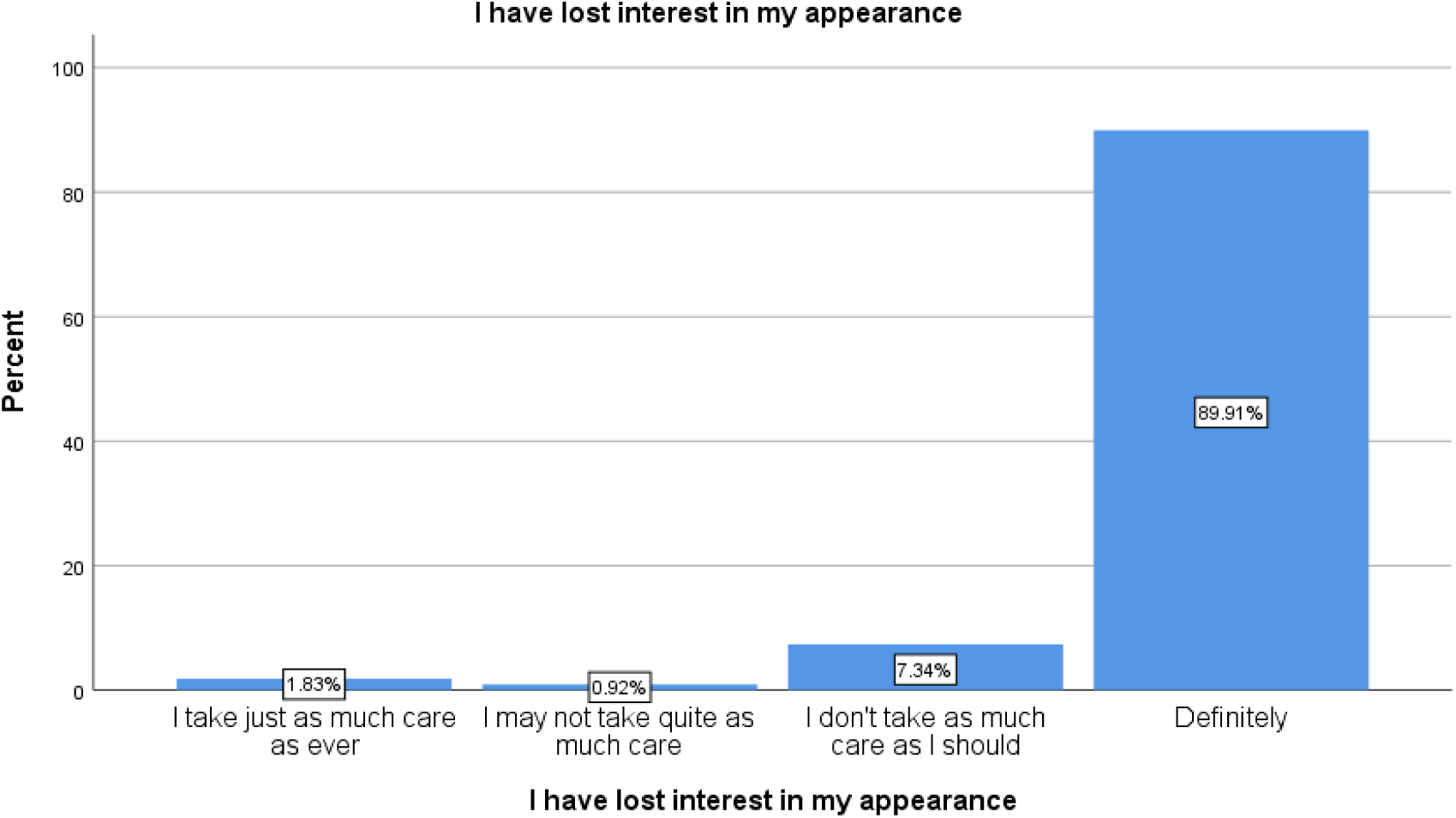

The above figure shows that the participants have lost interest in their appearance is definitely 98(89.9%), followed by I don’t take as much care as I should 8(7.3%), I take just as much care as ever 2(1.8%) and I may not take quite as much care 1(0.9%).

### TYPES OF STROKE

| Types of Strokes |  | Prevalence | Percent | Cumulative Percent |
| --- | --- | --- | --- | --- |
|  | Ischemic Stroke | 52 | 47.7 | 48.1 |
|  | Hemorrhagic Stroke | 56 | 51.4 | 100.0 |
|  | Total | 108 | 99.1 |  |
| System |  | 1 | .9 |  |
| Total |  | 109 | 100.0 |  |

The above table shows the type of stroke in the participants. The participants having hemorrhagic stroke 56(51.4%), followed by ischemic stroke 52(47.7%).

### DEPRESSION AMONG THE PARTICIPANTS

|  |  | Prevalence | Percent | Cumulative Percent |
| --- | --- | --- | --- | --- |
|  | 0-7 | 40 | 36.7 | 36.7 |
|  | 8-10 | 23 | 21.1 | 57.8 |
|  | 11-21 | 46 | 42.2 | 100.0 |
|  | Total | 109 | 100.0 |  |

The above table shows that the participants having depression ranging from 11-21 46(42.2%), followed by 0-7 40(36.7%) and 8-10 23(21.1%).

### ANXIETY AMONG THE PARTICIPANTS

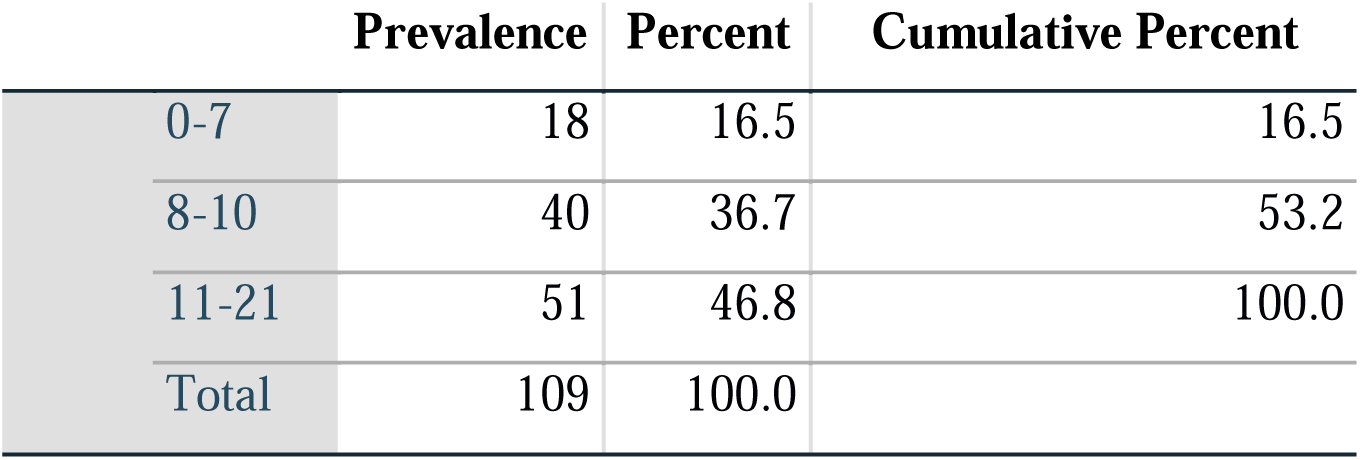

|  | Prevalence | Percent | Cumulative Percent |
| --- | --- | --- | --- |
| 0-7 | 18 | 16.5 | 16.5 |
| 8-10 | 40 | 36.7 | 53.2 |
| 11-21 | 51 | 46.8 | 100.0 |
| Total | 109 | 100.0 |  |

The above table shows that the participants having anxiety ranging from 11-21 51(46.8%), followed by 8-10 40(36.7%) and 0-7 18(16.5%).

## DISCUSSION

In the current study the prevalence of Post Stroke Depression score ranging from 0-7(36.70%), 8-10(21.10%) and 11-21(42.2%). The participant’s age ranged from 20-85 years old with mean age of 44.39 years and standard deviation of 14.748. Among 109 participants, depression was 63.3% and anxiety was 83.5%.The participants had no concern with depression was 36.70% and anxiety was 16.5%. The female participants had prevalence of PSD was 59(54.13%) and male participants had 50(45.87%), so female participants were more affected than male participants to concern with PSD.

FA I Marhiagbe, A Owolabi et al conducted a study in Africa in 2015 that reveals that PSD is a relatively common psychological condition occurring after a stroke and it only ranks behind anxiety and the incidence ranges from 5% to 20%;it was found to be as high as 60% in another related study. The following study was conducted in USA by Haresh M.Tharwani, MD et-al about recently advances in post-stroke depression in which prevalence varies from 20% to 80% it is under diagnosed and under treated (7,8).Much of the PSD research in the last year has focused on the neurochemical environment after stroke. St Paolucci et-al conducted a longitudinal study about The Italian multicenter observational study on post stroke depression (DESTRO). A total of 53 centers consequently admitted 1064 patients with ischemic or hemorrhagic stroke assessing the periodically in the first 9 months after the event. PSD was detected about 80% depression develop within 3 months of the stroke.

A study conducted in Egypt by Eman M.Khedr(1-2)et-al about PSD which is a developing country that’s why results of this study is slightly different from under developing countries in Egypt smoking and low economics condition is the major risk factors of PSD while another studies the major risk factors are anxiety depression. A study was conducted in Iran by Sahar Dalvand et-al about prevalence of PSD in Iran which is a systematic review and meta-analysis study which show that ratio of stroke in Iran is much high it means the sample size of this study is much large than our study.

## CONCLUSION

This study was performed among 109 research participants to find out the prevalence of post stroke depression among stroke patients admitted in Khyber teaching hospital Peshawar Hospital. In current study we have concluded a high prevalence of post stroke depression ranging from 65-74 years of age in Khyber teaching hospital Peshawar Hospital. The most affected were females as 59(54.1%) then males were 50(45.9%). We conclude that females suffering from stroke have more chances of post stroke depression.

## Data Availability

All data produced in the present work are contained in the manuscript

